# County-level factors influence the trajectory of Covid-19 incidence

**DOI:** 10.1101/2020.05.05.20092254

**Authors:** Ashley Wendell Kranjac, Dinko Kranjac

**Affiliations:** Department of Sociology, Chapman University, 1 University Drive, Orange, CA, 92866, USA; Department of Psychology, University of La Verne, 1950 Third Street, La Verne, CA, 91750, USA

**Keywords:** SARS-CoV-2, Covid-19, Community, Public Health Medicine

## Abstract

With new cases of Covid-19 surging in the United States, we need to better understand how the spread of novel coronavirus varies across all segments of the population. We use hierarchical exponential growth curve modeling techniques to examine whether community social and economic characteristics uniquely influence the incidence of Covid-19 cases in the urban built environment. We show that, as of May 3, 2020, confirmed coronavirus infections are concentrated along demographic and socioeconomic lines in New York City and surrounding areas, the epicenter of the Covid-19 pandemic in the United States. Furthermore, we see evidence that, after the onset of the pandemic, timely enactment of physical distancing measures such as school closures is imperative in order to limit the extent of the coronavirus spread in the population. Public health authorities must impose nonpharmaceutical measures early on in the pandemic and consider community-level factors that associate with a greater risk of viral transmission.

## Introduction

The presence of severe acute respiratory syndrome coronavirus 2 (SARS-CoV-2) was initially detected in Wuhan, China in December 2019 [1]. Since then, the outbreak of coronavirus disease 2019 (Covid-19), the illness caused by SARS-CoV-2, has spread worldwide [2]. As of May 3, 2020, the United States finds itself atop the list of countries most heavily impacted by the pandemic, both in terms of total diagnoses and fatalities [3]. Within the United States, the residents of the State of New York are disproportionately affected by SARS-CoV-2 virus, with New York City accounting for the vast majority of cases and deaths in the country [4]. Following the first confirmed New York case in March of 2020, the incidence of Covid-19 in and around New York City continues its alarming exponential rise [4]. The upward trend, however, is not uniformly patterned, as evidenced by substantial spatial heterogeneity of confirmed cases [5]. Higher population density increases the potential for virus transmission and may, in part, account for geographic differences in numbers of COVID-19 cases [6].

In addition to the readily predictable effects of population density, communities exhibit clear stratification along dimensions of demographic and socioeconomic characteristics that associate with vulnerability such as poverty that influences infectious disease patterning [6]. Indeed, social epidemiologists and medical sociologists argue that individuals and groups possessing fewer social, educational, and economic resources may prove less able to avoid areas where infectious disease runs rampant [7]. Here, we propose that variation in existing socioeconomic, demographic, and structural disparities may contribute to community-level differences in SARS-CoV-2 infection rates in New York City and surrounding areas.

Published reports indicate that community-level characteristics affect the incidence of numerous communicable diseases [7], but data specific to Covid-19 confirmed cases are currently inconsistent [8-12]. Notably, findings vary substantially by the modeling technique used, inclusion/exclusion of independent variables, granularity (e.g., county-level vs. state-level), and domain of outcome being measured (e.g., concentrated disadvantage vs. social vulnerability index). Consequently, this study adds to the existing literature, and helps clarify the impact of county-level demographic and socioeconomic disparities on Covid-19 incidence.

## Methods

In our analyses, spread of Covid-19 is captured by the number of daily confirmed cases per county extracted from USAFacts [13]. USAFacts confirms county-level data by directly referencing both state (New York State Department of Health) and local (New York City Department of Health) public health agencies [8]. Here, time is measured as the number of days since the first diagnosed case per county. We include a spatial lag comprised of the average geodetic distance between each county center to account for dependencies in Covid-19 confirmed cases across counties [14]. K–12 school closure dates were included to account for variation in time of enactment of physical distancing measures across counties since the first reported case in each of the counties [15-16]. County-level measures of socioeconomic factors are generated using 2014–2018 American Community Survey data, and include median age, population density, percent of Black residents, percent below the federal poverty level, percent receiving public assistance, percent of female-headed households, and percent unemployed [17]. We use an index of concentrated disadvantage and a measure of population density as socioeconomic indicators of counties. For concentrated disadvantage we followed Sampson *et al*. and take the first dimension of a principle components factor analysis on all the county-level characteristics described, with the exception of population density and median age [18].

We test the impact of distinct sociodemographic environments on the relative risk of Covid-19 infection using hierarchical exponential growth curve modeling techniques that account for correlation in the number of cases by county. All models use maximum likelihood estimation with adaptive quadrature that adjust for problems that would otherwise downwardly bias estimated standard errors [19]. Preliminary analyses indicate that time is most appropriately captured by a polynomial time function due to non-linearity, and thus we present only results from these models. The estimated coefficients in our models represent the estimated effect of concentrated disadvantage on change in the infection incidence rate over time. In the models, time points are nested within counties. The fully specified level-1 model fits the number of cases per day as a function of time across observations for each county, while simultaneously accounting for variation in the enactment of the physical distancing measures and a spatial lag. The fully specified level-2 model fits the level-1 intercept and coefficients across all counties as a function of the county-level characteristics.

## Results

Table 1 displays the number of confirmed Covid-19 cases per county, as well as estimated means and standard deviations of the covariates for each county. Queens County has the highest number of reported cases (n=53,640), followed by Kings County (n=46,839), whereas New York County has the lowest number of reported cases (n=22,741). Enactment of K–12 school closures began on March 16, which is 9 days after the first reported case in Bronx and Suffolk Counties, but 15 days after the first reported case in New York County. The median age of the population varies significantly across counties, from a high of 42 years old in Nassau County to a low of 34 years old in Bronx County. Population density varies substantially across counties, with Kings County the most concentrated (37,253). Relative to other counties, Kings County is one of the most disadvantaged, with the highest percentage of the population below poverty (17.1%), highest percentage of female-headed households (30%), and highest unemployment rate (6.3%). Bronx County has the highest percentage of Black residents (36.4%) and the highest percentage of the population receiving public assistance (8%).

**Table 1.** Means and Standard Deviations for Dependent and Independent Variables, New York Counties through May 3, 2020; N= 263,655.

|  |  |  | Bronx County |  | Kings County |  | Nassau County |  | New York County |  | Queens County |  | Suffolk County |  | Westchester County |  | Diff. |
| --- | --- | --- | --- | --- | --- | --- | --- | --- | --- | --- | --- | --- | --- | --- | --- | --- | --- |
| Independent Variables |  |  | Mean | Std.Dev. | Mean | Std.Dev. | Mean | Std.Dev. | Mean | Std.Dev. | Mean | Std.Dev. | Mean | Std.Dev. | Mean | Std.Dev. |  |
|  | Time |  |  |  |  |  |  |  |  |  |  |  |  |  |  |  |  |
|  | Time |  | 28.33 | 13.50 | 21.58 | 19.94 | 30.05 | 14.57 | 30.93 | 15.76 | 38.13 | 18.63 | 34.06 | 15.24 | 33.59 | 16.65 | *** |
|  | Time<sup>2</sup> |  | 982.63 | 826.42 | 858.56 | 1077.47 | 1112.51 | 933.18 | 1202.05 | 1064.52 | 1796.94 | 1258.70 | 1389.64 | 896.39 | 1370.69 | 997.89 | *** |
|  | Time<sup>3</sup> |  | 38409.49 | 46155.81 | 38941.00 | 58733.18 | 46314.12 | 54731.53 | 53129.67 | 65331.32 | 91218.24 | 91218.89 | 60497.03 | 47576.88 | 60905.41 | 59149.01 | *** |
|  | Spatial Lag |  |  |  |  |  |  |  |  |  |  |  |  |  |  |  |  |
|  | Distance in km |  | 29.95 | -- | 14.23 | -- | 18.89 | -- | 19.68 | -- | 31.34 | -- | 33.34 | -- | 106.42 | -- |  |
|  | Physical Distancing |  |  |  |  |  |  |  |  |  |  |  |  |  |  |  |  |
|  | Number of Days Since 1st Case |  | 9.00 | -- | 12.00 | -- | 12.00 | -- | 15.00 | -- | 10.00 | -- | 9.00 | -- | 14.00 | -- | *** |
|  | County Context |  |  |  |  |  |  |  |  |  |  |  |  |  |  |  |  |
|  | Median Age |  | 33.90 | 0.00 | 35.10 | 0.00 | 41.60 | 0.00 | 37.30 | 0.00 | 38.70 | 0.00 | 41.30 | 0.00 | 40.80 | 0.00 | *** |
|  | Population Density |  | 34194.00 | 0.00 | 37253.00 | 0.00 | 4763.00 | 0.00 | 72056.00 | 0.00 | 21132.00 | 0.00 | 1632.00 | 0.00 | 2250.00 | 0.00 | *** |
|  | Concentrated Disadvantage |  |  |  |  |  |  |  |  |  |  |  |  |  |  |  |  |
|  | % Black Residents |  | 36.40 | 0.00 | 33.90 | 0.00 | 13.10 | 0.00 | 17.10 | 0.00 | 19.70 | 0.00 | 8.90 | 0.00 | 16.20 | 0.00 | *** |
|  | % Below Poverty |  | 26.10 | 0.00 | 17.10 | 0.00 | 4.00 | 0.00 | 12.70 | 0.00 | 10.50 | 0.00 | 4.70 | 0.00 | 6.20 | 0.00 | *** |
|  | % Receiving Public Assistance |  | 7.90 | 0.00 | 4.80 | 0.00 | 1.30 | 0.00 | 2.80 | 0.00 | 3.20 | 0.00 | 1.80 | 0.00 | 1.80 | 0.00 | *** |
|  | % Female Headed- Households |  | 29.90 | 0.00 | 18.20 | 0.00 | 11.20 | 0.00 | 11.10 | 0.00 | 16.00 | 0.00 | 11.10 | 0.00 | 12.50 | 0.00 | *** |
|  | % Unemployed |  | 6.30 | 0.00 | 4.40 | 0.00 | 2.80 | 0.00 | 3.80 | 0.00 | 4.00 | 0.00 | 3.10 | 0.00 | 3.80 | 0.00 | *** |
| Dependent Variable |  |  |  |  |  |  |  |  |  |  |  |  |  |  |  |  |  |
|  | Number of Confirmed COVID-19 Cases |  | 38,916 |  | 46,839 |  | 36,780 |  | 22,741 |  | 53,640 |  | 34,855 |  | 29,884 |  | *** |

Source: Data are from USAFacts, NYC - The Official Website of the City of New York, Suffolk County Government, and 2014-2018 American Community Survey Data
*Note: Asterisks indicate significant difference evaluated using one-way MANOVA with county-level factors as the dependent variables and county as the independent variable*
\* $p < .05$ , \*\* $p < .01$ , \*\*\* $p < .001$

The results from our Poisson hierarchical polynomial growth regression models are shown in Table 2. For ease of interpretation, estimated fixed and random effects are displayed in terms of incidence rate ratios (IRR). Model 1 estimates the average incidence rate of Covid-19 cases across counties over time. Model 2 assesses IRR, with the spatial lag and physical distancing measures included at level-1. Model 3 adds the county-level characteristics of median age and concentrated disadvantage at level-2. We control for population density in all models.

**Table 2.** Poisson Hierarchical Polynomial Growth Regression Models Predicting Counts of Confirmed Covid-19 Cases through May 3, 2020, New York Counties; N=263,655.

|  |  | Model 1 |  |  | Model 2 |  |  | Model 3 |  |
| --- | --- | --- | --- | --- | --- | --- | --- | --- | --- |
|  |  | IRR | SE |  | IRR | SE |  | IRR | SE |
| Intercept |  | 1.05*** | 0.00 |  | 1.11*** | 0.04 |  | 1.19*** | 0.08 |
|  | Time | 1.68*** | 0.00 |  | 1.66** | 0.29 |  | 1.66** | 0.29 |
|  | Time<sup>2</sup> | 0.94*** | 0.00 |  | 0.99*** | 0.00 |  | 0.99*** | 0.02 |
|  | Time<sup>3</sup> | 1.00*** | 0.00 |  | 1.00*** | 0.00 |  | 1.00*** | 0.00 |
|  | Spatial Lag |  |  |  | 1.00 | 0.00 |  | 1.00 | 0.00 |
|  | Physical Distancing |  |  |  |  |  |  |  |  |
|  | K-12 School Closure |  |  |  | 0.21*** | 0.02 |  | 0.21*** | 0.02 |
| County Context |  |  |  |  |  |  |  |  |  |
|  | Median Age |  |  |  |  |  |  | 1.31* | 0.15 |
|  | Population Density | 1.00* | 0.00 |  | 1.00 | 0.00 |  | 1.00 | 0.00 |
|  | Concentrated Disadvantage |  |  |  |  |  |  | 2.74** | 1.03 |
| Random Effects |  |  |  |  |  |  |  |  |  |
|  | Intercept | 0.99*** | 0.00 |  | 0.88*** | 0.00 |  | 1.08*** | 0.00 |
|  | Time | 0.57*** | 0.00 |  | 0.51*** | 0.00 |  | 0.38*** | 0.01 |
|  | Time<sup>2</sup> | 0.64*** | 0.00 |  | 0.60*** | 0.00 |  | 0.35*** | 0.02 |
|  | Time<sup>3</sup> | 0.63*** | 0.00 |  | 0.58*** | 0.00 |  | 0.22*** | 0.00 |
|  | Concentrated Disadvantage | 0.18** | 0.08 |  | 0.29** | 0.03 |  | 0.41** | 0.10 |

*Source: Data are from USAFacts, NYC - The Official Website of the City of New York, Suffolk County Government, and 2014-2018 American Community Survey Data*
*\*p <.05, \*\*p <.01, \*\*\*p<.001*

From the onset of the outbreak, Covid-19 IRR averages around 1.05 (p<0.001) across counties, with an approximate 68% increase of cases per day (linear: 1.68, p<0.001; quadratic: 0.94, p<0.001; polynomial: 1.00, p<0.001.) Turning next to Model 2, in the early phase of the outbreak, the estimated average case incidence rate for counties, without the physical distancing measures enacted, and when holding population density and spatial differences constant, is 1.11 (p<0.001). The number of Covid-19 cases is rapidly increasing over time, evidenced by the positive values for the linear (1.66, p<0.001), negative values for the quadratic (0.99, p<0.001), and positive values for the polynomial (1.00, p<0.001) component of the time slope (see, Model 2 in Table 2). Model 2 further indicates that the enactment of physical distancing measures led to a 79% reduction in the daily number of reported cases (0.21, p<0.001).

Model 3 adds the county-level factors and indicates that as median age increases across counties, the daily number of cases increases by approximately 31% (p<0.05). After accounting for all the county-level characteristics, higher levels of concentrated disadvantage associate with a significant, nearly 3-fold, increase in the incidence rate of Covid-19 cases (2.74, p<0.01). As shown in the lower portion of Table 2, the random effects estimates indicate that the patterns of growth do indeed vary significantly across counties, both in terms of concentrated disadvantage (0.41, p<0.01), as well as in the initial levels (1.08, p<0.001), linear (0.38, p<0.001), quadratic (0.35, p<0.001), and polynomial (0.22, p<0.001) components of the incidence curve. Examples of this divergence are illustrated in Figure 1, where we graph the predicted curve for counties with low, medium, and high levels of concentrated disadvantage using the polynomial terms.

**Figure 1.**
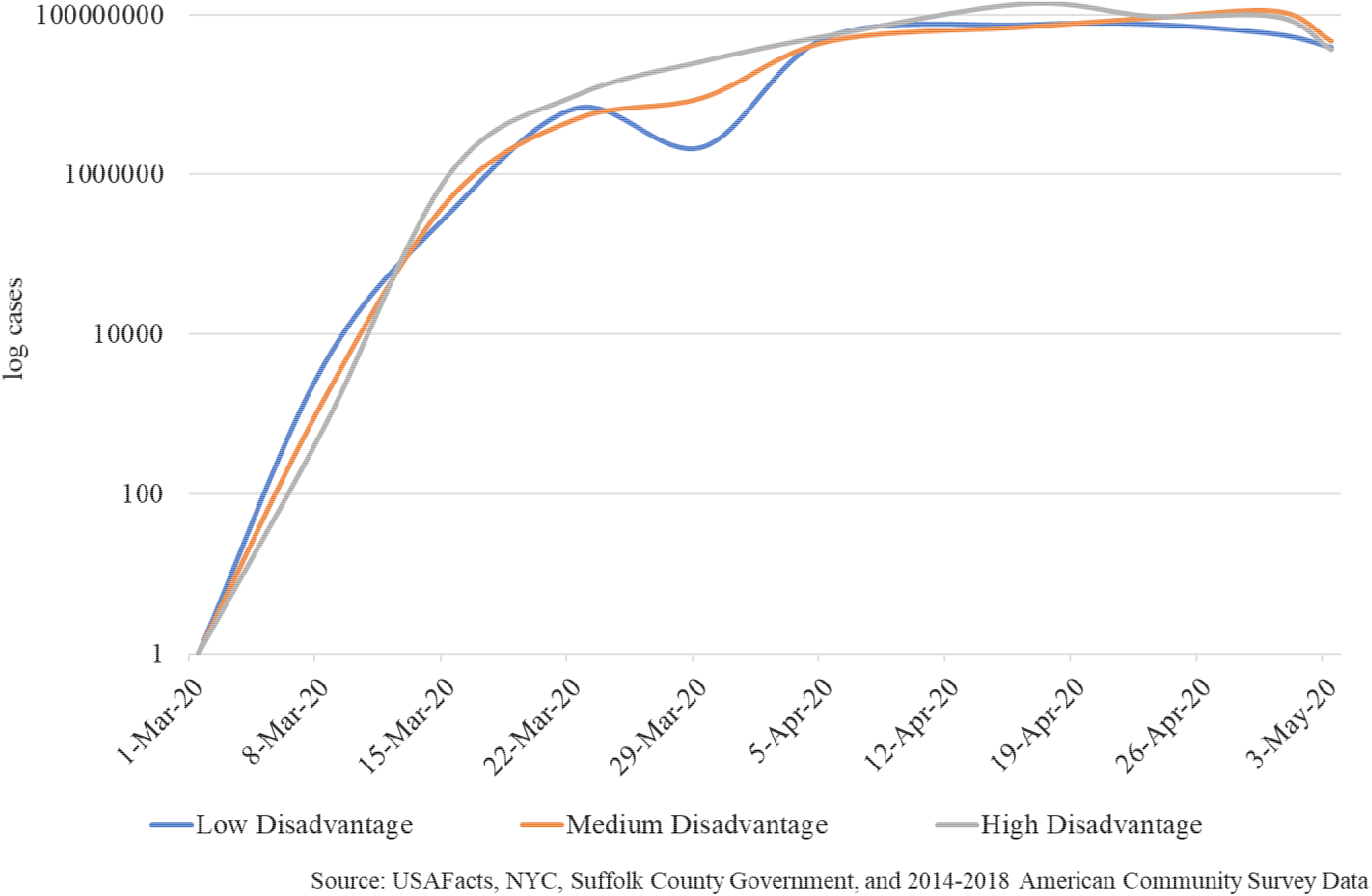
Covid-19 Growth Trajectories by Concentrated Disadvantage through May 3, 2020, N=263,655

## Discussion

Overall, our results through May 3, 2020, indicate that the number of documented Covid-19 infections varies widely both *between* counties and over time *within* counties. For all counties, we observe a substantial increase in the daily number of diagnoses over time and see evidence that earlier enactment of personal protective behaviors, here measured by K-12 school closure dates, likely altered the trajectory of the outbreak. This later finding clearly underscores the importance of timely introduction of mitigation measures to prevent virus diffusion, particularly when used in combination and enacted without delay [20]. Thus, in line with prior reports, the nonpharmaceutical interventions may help mitigate early spread of SARS-CoV-2 in urban areas with high population density [21].

Here, we add to this literature, and show that concentrated disadvantage is a potent ingredient in the observed pattern of detected Covid-19 infections across New York counties. We see evidence that the influence of communities on the spread of infectious disease reaches far beyond the physical characteristics of the urban built environment [22]. Furthermore, if community sociodemographics exhibit elevations in median age and/or have higher prevalence of disadvantage (e.g., poverty), such factors will likely have far-reaching consequences for SARS-CoV-2 infection incidence, and, Covid-19-related case fatality rates [23]. Although it is beyond the scope of our analysis to explain precisely why this variation exists, Covid-19 has exposed major social and economic disparities that already exist in the United States. Indeed, individual’s risk of being infected is higher if they reside in counties of New York with more socioeconomic disadvantage. This may be important because, at least at the county level, living in areas characterized by disadvantage increases the risk of infection.

To the best of the authors’ knowledge, this is the first study that employs hierarchical exponential growth curve modeling techniques to examine whether county-level demographic and socioeconomic characteristics influence the incidence of Covid-19 cases in New York City and surrounding areas. This study, however, is not without limitations. As this devastating pandemic continues to unfold in this epicenter of the outbreak in the United States, we assume that the number of Covid-19 confirmed cases will likely shift in response to improved testing and reporting practices, and thus, over time, enable the emergence of more accurate and useful data. Still, we use the latest data available to us [8]. Similarly, our sample is drawn from the New York metropolitan region, reducing the generalizability of our findings to a particular portion of people in the City of New York and surrounding areas, through May 3, 2020. Furthermore, in order to reduce virus transmission and keep case-fatality rates as low as possible, the New York State government instituted a number of mitigation efforts aside from school closures, including social distancing, school and workplace closures, cancellation of large-scale public gatherings, and stay-at-home orders [24]. In the present analysis, we use school closure dates to account for variation in time of enactment of physical distancing measures across counties. Thus, our inclusion of one variable is by no means comprehensive. However, if school closures were the only measure enacted, intervention effectiveness would be substantially reduced and variation in Covid-19 incidence across counties would be magnified [20-21]. Similarly, although we are unable to confirm whether individuals who tested positive for SARS-CoV-2 reside in the area where testing was performed, local government agencies direct people to contact their local health department for Covid-19-related concerns [24], and since stay-at-home measures were enacted at the same time as school closures we are confident in the results reported here.

Taken together, the dire circumstances in which we find ourselves demand better understanding of how distinctive geographic spaces influence infection incidence in order to potentially isolate the community-level factors that associate with higher likelihood of Covid-19 diagnosis and disease progression. Our analysis serves as a substantial jumping off point for future researchers interested in disentangling which community factors relate to SARS-CoV-2 transmission.

## Data Availability

Data are publicly available.

https://www.ecdc.europa.eu/en/publications-data/download-todays-data-geographic-distribution-covid-19-cases-worldwide

https://www.arcgis.com/home/webscene/viewer.html?layers=628578697fb24d8ea4c32fa0c5ae1843

https://usafacts.org/visualizations/coronavirus-covid-19-spread-map/

## Acknowledgements

The authors thank Robert L. Wagmiller for his conceptual and methodological help and Gary W. Boehm for his assistance in early drafts of the manuscript.

